# Early degeneration of motor pathways in prodromal Parkinson’s disease: A fixel-based structural connectivity analysis

**DOI:** 10.64898/2026.01.07.26343523

**Authors:** AmirHussein Abdolalizadeh, Stephanie Rosemann, Masoud Tahmasian, Christiane M. Thiel

## Abstract

**Background:** Idiopathic REM sleep behavior disorder (iRBD) and hyposmia are the strongest clinical indicators of prodromal Parkinson’s disease, reflecting distinct proposed pathways of α-synuclein pathology propagation. According to the body-first/brain-first model, iRBD represents a body-first phenotype with early brainstem involvement, whereas hyposmia may reflect a brain-first phenotype with early limbic involvement. We therefore hypothesized that these markers would show distinct fiber-specific white matter alterations, with iRBD and hyposmia primarily affecting brainstem and limbic pathways, respectively.

**Methods:** Diffusion and structural MRI from prodromal participants in the Parkinson’s Progression Markers Initiative (PPMI; n = 88) were analyzed using fixel-based analysis, which quantifies macro- and microstructural fiber-specific measures of fiber density, fiber cross-section, and fiber density-cross-section. We assessed the association between these measures and RBD severity, olfactory performance, and motor symptom severity. In a subset with polysomnography data (n = 37), we compared fiber-specific metrics between polysomnography-confirmed and polysomnography-negative iRBD. Voxel-based morphometry was also conducted to assess grey matter volume correlates of prodromal symptom severity.

**Results:** Greater RBD severity was associated with reduced fiber cross-section in the pons, right internal capsule, and left temporal lobe, corresponding to the right corticospinal tract and left inferior longitudinal fasciculus. The lower fiber cross-section in the latter was associated with vivid dreams. Polysomnography-confirmed iRBD showed lower fiber density in the midbrain and bilateral internal capsules, corresponding to left corticospinal and right frontopontine tracts. Lower corticospinal fiber density predicted worse fine motor performance. No diffusion or grey matter correlates emerged for hyposmia.

**Conclusions:** Prodromal Parkinson’s disease shows selective degeneration of corticospinal, frontopontine, and inferior longitudinal fasciculus pathways that scale with iRBD severity, showing early brainstem and motor pathways involvement in body-first phenotypes. The association between reduced corticospinal fiber density and fine motor impairment demonstrates that these tract-level alterations are not only detectable in the prodromal phase but are already clinically meaningful. No detectable diffusion or grey matter correlates were observed for hyposmia.

These findings identify iRBD-specific white matter vulnerability in the prodromal phase and highlight fiber-specific diffusion metrics as sensitive markers of early Parkinson’s disease-related change.

## 1. Background

Parkinson’s disease is the fastest-growing neurodegenerative disorder in terms of prevalence, affecting more than 8.5 million people worldwide, with 25.2 million people expected to live with the disease by 2050.^1^ It is defined by its cardinal motor manifestations, including bradykinesia, rigidity, and rest tremor; however, it is preceded by a largely understudied prodromal phase that can last 10-20 years. Two non-motor markers of the prodromal phase, namely Idiopathic Rapid Eye Movement Sleep Behavior Disorder (iRBD) and hyposmia (i.e., olfactory loss), are the strongest clinical harbingers of Parkinson’s disease, each conferring a several-fold increased risk of developing the disease.^2,3^ Although these symptoms provide a unique window into the early disease process, the neurobiological mechanisms underlying their emergence and their relationship to later neurodegeneration remain poorly understood, limiting progress in early diagnosis and the potential development of disease-modifying treatments.

Current research models classify Parkinson’s disease based on the origin of α-synuclein pathology into “body-first” and “brain-first” clinicopathological phenotypes.^2^ In the body-first phenotype, α-synuclein aggregates are hypothesized to originate in the peripheral autonomic nervous system, particularly the gastrointestinal system, and to propagate centrally via the vagus nerve to the dorsal motor nucleus and pontine structures. In contrast, the brain-first phenotype is hypothesized to begin in the olfactory bulb, with subsequent propagation through the limbic system toward the brainstem.^4,5^ Given that iRBD often precedes motor symptoms by many years and is closely linked to autonomic dysfunction, it is generally considered a clinical marker of a body-first phenotype, whereas isolated hyposmia aligns more with a brain-first origin. The body-first/brain-first framework, therefore, predicts early structural changes in pontine and limbic white matter pathways, aligning with prodromal markers of iRBD and hyposmia, respectively.^2,6^ Because such tract-level changes likely emerge before overt dopaminergic dysfunction or grey matter atrophy,^7,8^ diffusion MRI provides an appropriate method to detect these early alterations in vivo.

Prior diffusion MRI studies using tensor-based metrics have shown reduced fractional anisotropy and increased mean diffusivity in the substantia nigra, internal and external capsules, midbrain, and pontine regions in patients with iRBD,^9,10^ as well as tractography evidence for alterations in the corticospinal tract, the anterior thalamic radiation, and the inferior fronto-occipital tract.^11,12^ However, conventional diffusion tensor metrics, such as fractional anisotropy and mean diffusivity, model each voxel with a single scalar value, obscuring tract-specific pathology in the many white matter regions that contain crossing fibers.^13^ Fixel-based analysis resolves this limitation by modelling individual fiber populations within each voxel (“fixels”). It allows the quantification of fiber density (FD), fiber cross-section (FC), and their product, fiber density-cross-section (FDC), separately.^14,15^ FD is proportional to intra-axonal volume within a fiber bundle and therefore reflects microstructural changes such as axonal loss or reduced packing. FC quantifies macrostructural alterations and shows changes in the cross-sectional size of a fiber bundle. Collectively, these fiber-specific metrics provide a more biologically specific and axon-oriented characterization of white matter architecture, overcoming key limitations of conventional tensor metrics.^15^

Fixel-based analysis has already been successfully applied in clinical Parkinson’s disease, revealing widespread white matter degeneration that progresses over time and correlates with greater clinical disability.^16–19^ Hence, investigating the prodromal stage with a fixel-based approach offers a unique opportunity to identify early, fiber-specific white matter changes that may underlie the later emergence of motor and cognitive symptoms in Parkinson’s disease.

In the present study, we examined the prodromal cohort of the Parkinson’s Progression Markers Initiative (PPMI) to test whether pathway-specific white matter changes associated with iRBD and hyposmia are already detectable before clinical diagnosis. To achieve fiber-resolved sensitivity to early micro- and macrostructural changes, we applied fixel-based analysis to diffusion MRI data and quantified FD, FC, and FDC. We further evaluated whether these fiber-specific measures showed associations with motor symptoms, and we complemented the white matter analysis with voxel-based morphometry to assess whether potential early changes are also evident in grey matter. Based on the body-first/brain-first hypothesis, we hypothesized that iRBD would show pronounced white matter structural degeneration in the brainstem, while hyposmia would mainly involve limbic pathways, yielding distinct fiber-specific patterns relevant to each prodromal marker.

## 2. Methods

### 2.1. Participants

We included all participants from the prodromal cohort of the Parkinson’s Progression Disease Initiative (PPMI; https://ppmi-info.org)^20^ dataset who had diffusion MRI acquired with high b-value recommended for fixel-based analysis (see *Image Acquisition*), resulting in a final sample of 88 individuals (Mean age + SD = 69.02 + 5.85, Sex = 49 females, 39 males).

According to the PPMI protocol, participants were included in the prodromal cohort following an abnormal dopamine transporter scan (DaTScan), and if they met the PPMI’s centrally applied “predictive-eligibility” algorithm, which requires hyposmia confirmed based on the University of Pennsylvania Smell Identification Test (UPSIT), an age ≥ 60 years (≥ 40 years for genetic-risk enrollees), and at least one additional Parkinson’s disease risk factor (e.g., clinician-identified iRBD, first-degree family history, or a recognised Parkinson’s disease-linked variant). The clinical diagnosis of iRBD was derived from the variable RBDDIAG (“Does the participant have a clinical diagnosis of RBD?”). Polysomnography status was taken from the variable RBDPSG (“Does the participant have a polysomnography consistent with diagnosis of RBD?”), with codes of 1, 0, and blank. For the polysomnography-related analysis, we analyzed a subset of 37 participants with a clinical iRBD diagnosis (RBDDIAG = 1) and available polysomnography data (RBDPSG ≠ blank), either consistent (RBDPSG = 1, n = 25) or inconsistent (RBDPSG = 0, n = 12) with the iRBD diagnosis. Although the prodromal PPMI cohort also includes individuals with genetic risk factors for Parkinson’s disease, none of the selected participants carried known disease-related genetic variants. The PPMI study protocol and ethics application were reviewed and approved by the IRBs of all participating sites, and every participant provided written informed consent prior to enrollment. Further details on the inclusion and exclusion criteria are available on the PPMI website (https://www.ppmi-info.org/study-design/research-documents-and-sops).

### 2.2. Clinical Evaluations

iRBD severity was assessed using the RBD Screening Questionnaire (RBDSQ). This self-reported questionnaire consists of 13 items grouped into 10 domains that evaluate various RBD-related clinical features, such as vivid dreams, aggressive dream content, and motor enactment behaviors. Each question can be answered with a yes or no, and the total score is calculated as the sum of the “yes” responses.^21^ Olfactory dysfunction was assessed using the 40-item UPSIT. Participants were presented with 40 different odors and asked to identify them correctly, with the total score reflecting the number of correct responses.^22^ Motor symptom severity was evaluated using the Movement Disorder Society – Unified Parkinson’s Disease Rating Scale (MDS-UPDRS) Part 3 total score, as well as subscale scores for bradykinesia, rigidity, and resting tremor (see Supplementary Table 1).^23^ Additionally, we used the short-form fine motor assessment scores from Neuro-QoL.^24^ This questionnaire assesses a patient’s ability to perform fine motor tasks, such as picking up a coin or writing with a pen, scoring each task from 1 (unable to do) to 5 (performing the task without any difficulty). Originally, higher scores indicate better performance, but for consistency in interpretation, we inverted the scale so that a higher score now reflects greater fine motor disability.

### 2.3. Image Acquisition

All images were acquired on 3T Siemens MRI scanners (Prisma and Skyra). PPMI is a multi-site dataset, and the protocol details (such as TR/TE) may vary slightly across acquisition sites. Each subject underwent a structural T1 scan acquired with a 1 mm isotropic voxel size. TR and TI for all T1 acquisitions were 2300 and 900 ms; however, TE varied between 2.98 ms (for most subjects) and 3.06 ms, with a flip angle of 8° and 9°. Fixel-based analysis is most sensitive when diffusion MRI is acquired with high b-values and sufficient angular resolution, which improves the reliability of fiber-specific metrics and the ability to resolve crossing fibers.^15^ Following these methodological considerations, we included only prodromal subjects who underwent the highest-quality diffusion MRI protocol in PPMI. This consisted of multi-shell diffusion MRI with b-values of b=700 s/mm^2^, b=1000 s/mm^2^, and b=2000 s/mm^2^, each sampled with 64 directions (with six non-diffusion images), voxel-size = 2 mm isotropic, phase-encoding direction = PA, multiband acceleration factor = 3, with an additional two non-diffusion volumes in reverse phase encoding direction. TR/TE was 3800/90 ms for 81 subjects and 3800/100 ms for seven subjects. For a detailed explanation of acquisition protocols, please refer to the PPMI MRI acquisition manual, available in their research documents: https://www.ppmi-info.org/study-design/research-documents-and-sops.

### 2.4. Fixel-based Analysis

We analyzed diffusion MRI data using MRtrix (version 3.0.4).^25^ The raw diffusion MRI data were first denoised,^26^ and the Gibbs ringing artifact was removed.^27^ Next, *dwifslpreproc,* utilizing FSL’s eddy (version 6.0.7.3), was used to apply distortion, eddy-current, and subject motion correction.^28–30^ Additionally, Eddy’s outlier detection and replacement tool^31^ and within-volume movement correction were also applied.^32^ Then, B1 field inhomogeneity correction with the N4 algorithm was done,^33^ and the preprocessed diffusion data were reoriented to the FSL standard orientation. To minimize the effect of extra-axonal signal in our fixel-based analysis,^34^ we extracted only b-value shells of 0 and 2000 s/mm^2^ for the following steps.

The *dhollander* algorithm was used to estimate tissue response functions for white matter, grey matter, and cerebrospinal fluid.^35,36^ A study-average response function was calculated for each tissue type. The diffusion data were upsampled to a voxel size of 1.25 mm, and a robust brain mask was generated based on the upsampled non-diffusion images using SynthSeg.^37,38^ SynthSeg was also applied to the T1 scans to extract robust total intracranial volumes (ICV), which were necessary for statistical analysis. Single-shell 3-Tissue Constrained Spherical Deconvolution (SS3T-CSD) was performed using the brain masks and the study-average response functions to obtain fiber orientation distributions (FODs) with MRtrix3Tissue (https://3tissue.github.io/),^39^ a fork of MRtrix3.^25^ FODs were normalized using log-domain intensity normalization.^40,41^ We then created a population-specific FOD template using FOD images of all subjects and registered individual FOD images to the group template.

We registered individual brain masks to the group FOD template using the warps made in the previous step, and built a study-specific voxel-wise template mask from the intersection of warped brain masks. Then, a fixel-wise analysis mask was generated within this voxel-wise template mask. FODs of subjects were segmented into fixels^42^ and reoriented to the template space. We calculated FD, FC, and FDC for each fixel. FD is calculated as the amplitude of FOD along a particular orientation and is proportional to the intra-axonal volume along the corresponding direction.^43^ FC is defined based on the warp required to transform fixels to the group template, and FDC is defined as the product of FC and FD.^14^ FC was log-transformed to account for its non-normal distribution (logFC).

Whole-brain tractography with 20 million fibers was then performed using the study-specific FOD template and brain mask with an angle threshold of 22.5, and minimum-maximum length of 10-250 mm.^44^ Spherical-deconvolution informed filtering of tractograms (SIFT) was then applied to the whole-brain tractography to retain 2 million fibers by removing false-positive tracts.^42^ Fixel-to-fixel connectivity was created based on the filtered tractogram, which was then used to smooth FD, logFC, and FDC only within that fixel’s fiber population.^25^

### 2.5. Selective Tractography with TractSeg

Fixels showing statistical significance in fixel-based analysis were mapped to their corresponding white matter tracts based on their anatomical location and orientation. We used TractSeg (version 2.8; https://github.com/MIC-DKFZ/TractSeg),^45–47^ a deep learning-based automated tractography tool, to perform selective tractography of those fiber tracts with 10000 streamlines using the three primary spherical harmonic peaks of the template FOD image. Then, the fixel density map of each tract was extracted and binarized to generate tract fixel masks. These masks were applied to the transformed fixel maps of the subjects to calculate subject-specific tract-average FD, logFC, and FDC.

To investigate RBDSQ subscales associated with tract-average fixel-metrics, we used an ANCOVA model in Rstudio^48^ using R version 4.2.2^49^ with tract-average fixel metrics as independent and yes/no responses of the subject in eight questions from RBDSQ, specifically assessing dream content (Questions 1-4) or motor enactment (6.1 - 6.4) as dependent variables.

To assess the association between tract fixel metrics and motor severity, we implemented a zero-inflated negative binomial model using *the zeroinfl* function from the *pscl* package in R.^50,51^ Zero-inflated models are best suited for scenarios with count data (symptom severity) that show a high frequency of zeros (i.e., patients not exhibiting motor symptoms during the early stages of the disease course). We chose zero-inflated over similar *hurdle* models,^52^ based on how they model zeros in our sample. In zero-inflated models, zeros have two origins: “sampling” (a subject not showing any symptoms) and “structural” (a subject cannot show any symptoms since it is not affected). However, hurdle models assume that all zero data have a structural origin only.^53^ Our sample includes a heterogeneous sample of prodromal Parkinson’s disease subjects, of which some are expected not to show any progression towards Parkinson’s disease, hence not manifesting the motor symptoms, making zero-inflated models better suited for this scenario. Age, gender, and logICV (for FC and FDC) were included as covariates in the model. P-values were adjusted by Benjamini-Hochberg correction.^54^

### 2.6. Voxel-Based Morphometry

We used CAT12 (https://neuro-jena.github.io/cat/)^55^ implemented in SPM12 (Statistical Parametric Mapping, Institute of Neurology, London, UK), for voxel-based morphometry (VBM) analysis. Structural scans were preprocessed using the default CAT12 pipeline (*segment*), which includes denoising, bias-field correction, and segmentation of the data into white matter, grey matter, and cerebrospinal fluid.^56^ The data were then registered to standard space using Diffeomorphic Anatomic Registration Through Exponentiated Lie Algebra (DARTEL).^57^ Then, a Gaussian kernel with 6 mm full-width at half maximum (FWHM) was applied to smooth the grey matter volume data, preparing the volumetric data for the statistical analysis.

Then, a general linear model within SPM was designed to assess morphological changes associated with RBDSQ, UPSIT, and polysomnography status. Age, sex, and total intracranial volume were entered as covariates.

## 3. Results

### 3.1. Patient Demographics

Patients’ demographics, and their RBDSQ, UPSIT, and motor scores are presented in Table 1. The distribution of motor severity scores is presented in Supplementary Figure 1.

**Table 1.** Subject demographics and evaluations. Variables that did not follow a normal distribution are marked with ^✝^and are reported as median [IQR1, IQR3]. Motor evaluations are reported both as median [IQR1, IQR3] and mean (SD), due to the presence of many zero values (zero-inflated). The percentage of zero incidences is added as a percentage. Scores on the fine motor scale were inverted, with higher values indicating worse fine motor performance. RBD: REM sleep behavior disorder, UPSIT: University of Pennsylvania Smell Identification Test, MDS-UPDRS: Movement Disorder Society - Unified Parkinson’s Disease Rating Scale.

| Demographics |  |
| --- | --- |
| Age (mean (SD)) | 69.02 (5.85) |
| Sex (F/M) | 49/39 |
| Prodromal Features |  |
| RBD Severity Questionnaire† | 5 [2 - 11] |
| UPSIT<sup>†</sup> | 20 [15 - 27.5] |
| **Motor Evaluations** |  |
| MDS-UPDRS Part 3 Total<br>Zeros (%) | 2 [0 - 7]; 4.05 (5.55)<br>36.90% |
| MDS-UPDRS Rigidity Subscale<br>Zeros (%) | 0 [0 - 0]; 0.41 (1.2)<br>82.76% |
| MDS-UPDRS Bradykinesia Subscale<br>Zeros (%) | 0 [0 - 3]; 2 (3.33)<br>53.49% |
| MDS-UPDRS Rest Tremor Subscale<br>Zeros (%) | 0 [0 - 0]; 0.14 (0.57)<br>94.32% |
| Neuro QoL Fine Motor<br>Zeros (%) | 0 [0 - 0]; 0.57 (1.98)<br>78.41% |

### 3.2. Fixel-Correlates of Prodromal Symptoms

To test our primary hypothesis regarding the two prodromal phenotypes, we performed a whole-brain fixel-based analysis examining associations between fixel metrics (FD, FC, and FDC) and prodromal symptoms, specifically UPSIT scores, and iRBD severity, as assessed by the RBDSQ. Greater iRBD severity was associated with reduced FC in the left temporal white matter, right internal capsule, and brainstem (Figure 1). No significant associations were found between UPSIT scores and any fixel metrics, suggesting that olfactory dysfunction in our sample did not exhibit detectable white matter microstructural correlates.

**Figure 1.**
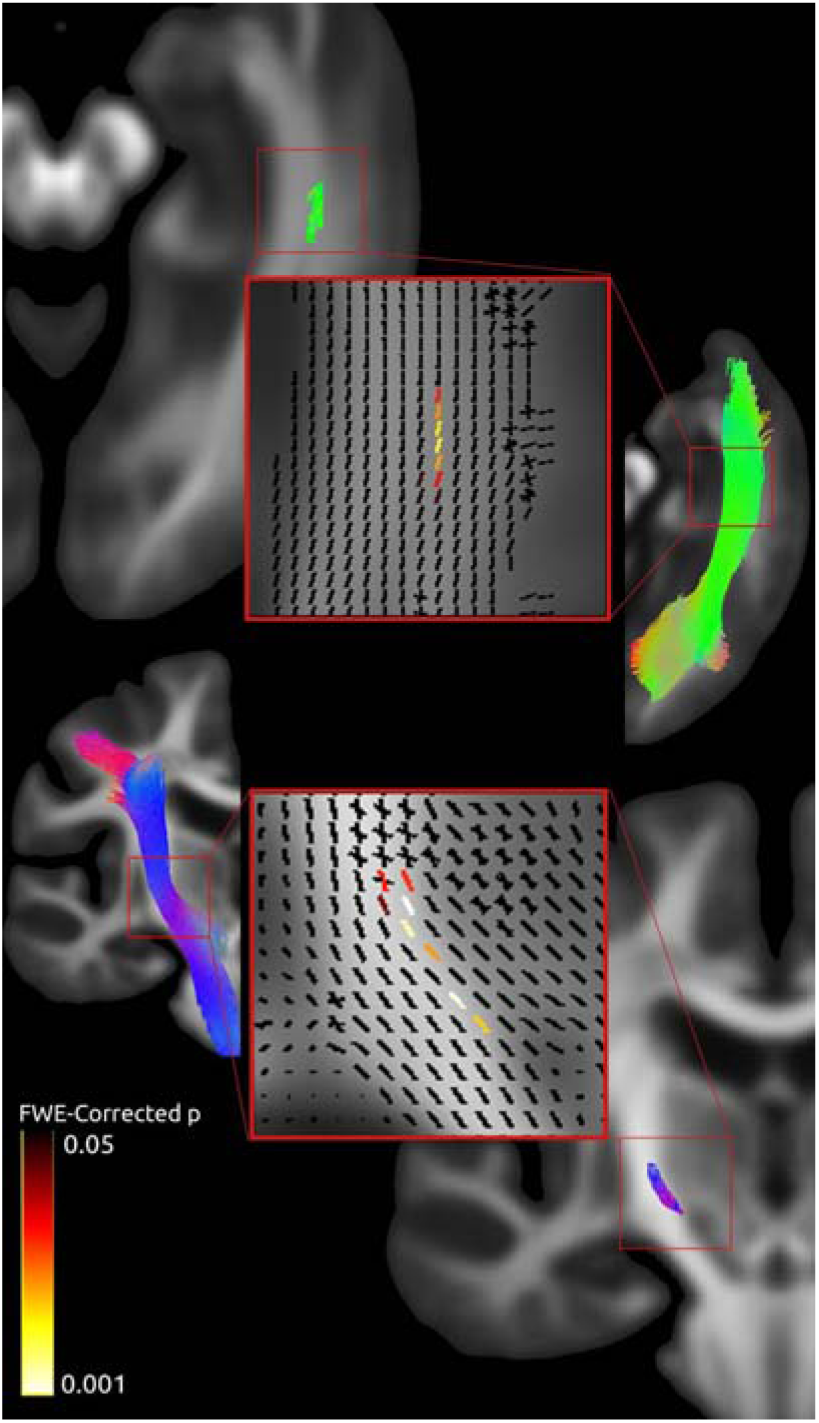
iRBD severity is associated with a lower cross-section of fibers corresponding to the corticospinal tract and inferior longitudinal fasciculus. Fixel-based analysis results associating fixel metrics with the severity of iRBD symptoms showed a reduced fiber cross-section in regions including the left temporal white matter (top) and internal capsule (bottom). There were significant regions in the brainstem (not shown; see Supplementary Figure 2). All these regions were mapped to the left inferior longitudinal fasciculus (top-right) and right corticospinal tract (bottom-left) based on their anatomical location and fiber orientation (see text). A close look at the fixel-based analysis results (red rectangles) reveals the significant fixels (hot-color-coded by p-values; FWE-corrected p<0.05) displayed within surrounding fiber communities in different directions (black fixels). The fibers are color-coded, blue/purple shows fibers elongated in the superior-inferior axis, and green shows fibers in the anterior-posterior axis. Tracts were generated by TractSeg.

To determine the white matter tracts corresponding to significant fixels associated with iRBD severity, we quantified their overlap with tracts generated by TractSeg (Supplementary Table 2). However, because TractSeg-generated tracts are not exclusive, we incorporated neuroanatomical knowledge to refine tract identification. The significant fixels in the left temporal white matter were aligned inferolaterally along the anterior-posterior axis, suggesting the left inferior longitudinal fasciculus as the most probable tract (Figure 1). The remaining significant clusters were located in the posterior part of the right internal capsule and the middle of the medulla (right below the pons) in axial slices, aligned along the superior-inferior axis, making the right corticospinal tract the most likely tract (Figure 1, Supplementary Figure 2).

Together, these results indicate selective macrostructural involvement of visuolimbic pathways, reflected by reduced FC in the left inferior longitudinal fasciculus and descending motor pathways, reflected by reduced FC in the right corticospinal tract, in association with iRBD severity. In contrast, hyposmia did not show detectable fiber-specific correlates.

### 3.3. Dream Content and Motor Enactment Correlates in RBD Severity

Given that the iRBD-related fixels localized to the left inferior longitudinal fasciculus, a tract linking visual and limbic regions and also implicated in dream recall frequency,^58,59^ we conducted a post hoc analysis to identify which RBD questionnaire items contributed most strongly to this association. We therefore examined how FC of the right inferior longitudinal fasciculus and left corticospinal tract related to items of the RBD severity questionnaire assessing dream content and motor enactment. Following multiple comparison correction, we found that the presence of vivid dreams (question 1: “I sometimes have very vivid dreams”) was significantly associated with lower FC in the inferior longitudinal fasciculus (*F*(1, 83) = 9.57, corrected *p* = 0.04, η^2^ = 0.104). This suggests that vivid dream presence accounted for 10.4% of the variance in the FC of the inferior longitudinal fasciculus in our sample. No other associations remained significant after multiple-comparison corrections (Table 2).

**Table 2.** Group mean (SD) for fiber cross-sections of the right corticospinal tract and the left inferior longitudinal fasciculus for the sub-items of dream content and motor enactment from the REM sleep behavior disorder screening questionnaire. The statistical test for comparison was an ANCOVA model with the fiber cross-section of the right corticospinal tract or left inferior longitudinal fasciculus as the outcome variables, and responses to dream content and motor enactment questions (yes/no) as the primary predictor. Age, sex, and intracranial volume were inserted as covariates.

|  | Right Corticospinal Tract |  |  | Left Inferior Longitudinal Fasciculus |  |  |
| --- | --- | --- | --- | --- | --- | --- |
| Questions | “Yes” | “No” | FDR corrected $p$-value | “Yes” | “No” | FDR corrected $p$-value |
| “I sometimes have very vivid dreams.” | 1.031<br>(0.084) | 1.04<br>(0.072) | 0.89 | 1.159<br>(0.092) | 1.201<br>(0.113) | **0.04\*** |
| “My dreams frequently have an aggressive or action-packed content.” | 1.028<br>(0.082) | 1.04<br>(0.08) | 0.18 | 1.166<br>(0.101) | 1.177<br>(0.099) | 0.07 |
| “The dream contents mostly match my nocturnal behaviour.” | 1.038<br>(0.083) | 1.031<br>(0.079) | 0.55 | 1.186<br>(0.089) | 1.161<br>(0.106) | 0.89 |
| "I know that my arms or legs move when I sleep." | 1.028<br>(0.079) | 1.041<br>(0.083) | 0.09 | 1.175<br>(0.098) | 1.167<br>(0.102) | 0.21 |
| "I have or had the following phenomena during my dreams: |  |  |  |  |  |  |
| Speaking, shouting, swearing, laughing loudly | 1.026<br>(0.08) | 1.041<br>(0.081) | 0.10 | 1.166<br>(0.104) | 1.176<br>(0.096) | 0.07 |
| Sudden limb movements, "fights" | 1.023<br>(0.082) | 1.044<br>(0.079) | 0.07 | 1.168<br>(0.104) | 1.175<br>(0.096) | 0.07 |
| Gestures, complex movements, that are useless during sleep, e.g., to wave, to salute, to frighten mosquitoes, falls off the bed | 1.026<br>(0.084) | 1.039<br>(0.078) | 0.10 | 1.167<br>(0.11) | 1.174<br>(0.092) | 0.07 |
| Things that fell down around the bed, e.g., bedside lamp, book, glasses" | 1.028<br>(0.091) | 1.036<br>(0.077) | 0.07 | 1.188<br>(0.106) | 1.165<br>(0.097) | 0.29 |

### 3.4. Polysomnography-confirmed iRBD is Associated with Lower Fiber Density in the Corticospinal and Frontopontine Tracts

As a follow-up validation analysis, we examined whether the white matter alterations observed for questionnaire-based iRBD severity would also emerge when testing for differences between polysomnography-confirmed and polysomnography-negative individuals, the clinical gold standard for diagnosis. Fixel-based analysis in this subset of patients showed lower FD in the posterior limb of the left internal capsule, anterior limb of the right anterior capsule, and cerebral peduncles as compared to patients with negative polysomnography findings (Figure 2). The cluster of significant fixels on the left side was continuous, extending in the superior-inferior axis from the posterior limb of the internal capsule to the middle third of the cerebral peduncle on the left side, suggesting the left corticospinal tract to be the most probable tract. On the right side, the significant fixels were situated in the anterior part of the internal capsule and more medially in the cerebral peduncle, indicating that the right frontopontine tract is the most probable candidate.

**Figure 2.**
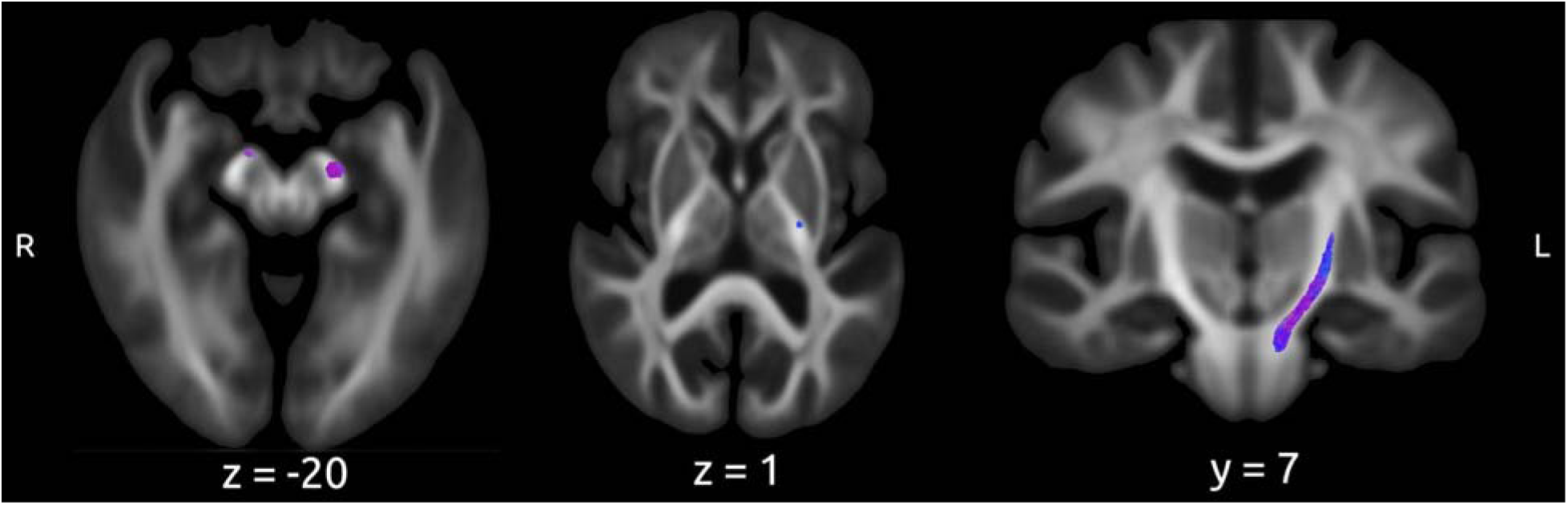
Patients with polysomnography-confirmed iRBD have lower fiber density in several white matter regions. Whole-brain fixel-based analysis comparing fixel metrics in the subset of subjects (n = 37) that had polysomnography data available showed lower fiber density in several areas in polysomnography-positive compared to polysomnography-negative subjects, including the anterior part of the right internal capsule, the posterior part of the left internal capsule, and the cerebral peduncles. The significant fixels anatomically correspond to the left corticospinal tract and right frontopontine tracts, given their location and trajectory.

In sum, these results indicate microstructural involvement of descending motor pathways in polysomnography-confirmed iRBD, with reduced fiber density in the left corticospinal and right frontopontine tracts marking early brainstem- and capsule-level degeneration.

### 3.5. Fiber Density of Corticospinal Tract Correlates with Fine Motor Problems

To assess the motor correlates of the tract-specific measures identified in the previous steps, we related quantitative motor evaluations (MDS-UPDRS part 3 total score, subscale scores of rigidity, bradykinesia, rest tremor, and fine motor) to tract-average fixel metrics. Using a negative binomial zero-inflated model and after adjusting for multiple comparisons, we found that lower FD in the left corticospinal tract was significantly associated with greater fine motor impairment (estimate (std. error) = -36.80 (11.66), corrected *p* = 0.03). In other words, these findings show that microstructural degeneration of the left corticospinal tract, identified in the polysomnography analysis, is already behaviorally relevant in the prodromal phase, as reflected by its association with fine motor impairment.

### 3.6. Voxel-Based Morphometry

To complement the fixel-based results and evaluate whether macrostructural grey matter changes accompany the observed white matter alterations, we investigated the volumetric relationships between grey matter and iRBD severity, hyposmia, and polysomnography status in all subjects using voxel-based morphometry. No grey matter differences survived family-wise error correction for any contrast involving iRBD severity, UPSIT scores, or polysomnography status. A small uncorrected cluster (300 voxels) was found in the inferior part of the right temporal lobe (Harvard-Oxford atlas: right temporal fusiform gyrus; Peak MNI coordinates 32 -4 -44), which was lower in polysomnography-confirmed iRBD compared to polysomnography-negative patients (uncorrected *p* = 0.007, FWE-corrected *p* = 0.079; Supplementary Figure 3). Hence, we did not observe grey matter alterations that covaried with prodromal symptoms.

## 4. Discussion

This study applied whole-brain fixel-based analysis in a mixed prodromal Parkinson’s disease population to characterize fiber-specific white matter alterations associated with the prodromal symptoms of iRBD and hyposmia. More severe iRBD symptoms were associated with lower FC in the right corticospinal tract and the left inferior longitudinal fasciculus, with the latter being explicitly affected in patients reporting the presence of vivid dreams. Polysomnography-confirmed iRBD was associated with lower FD in two major descending motor pathways, the left corticospinal and right frontopontine tracts. Across the entire cohort, lower FD in the left corticospinal tract was associated with worse fine motor function, providing a behavioral correlate of this early pathway involvement. Contrary to our expectation, we did not observe fiber-specific alterations related to olfactory performance, nor grey matter correlates of either prodromal marker. Together, these findings indicate that early white matter alterations in descending motor and visuolimbic pathways are specifically associated with iRBD and may precede the onset of overt motor signs in prodromal Parkinson’s disease.

These results partly align with our hypotheses, revealing early involvement of brainstem-traversing motor pathways in association with iRBD, whereas the anticipated limbic correlates of hyposmia were not detected. Although the observed alterations were located in pontine and midbrain-traversing sections of the descending motor tracts rather than in classically defined pontine nuclei, their trajectories pass through brainstem regions proposed to be affected early in body-first Parkinson’s disease.^2^ This pattern, therefore, remains consistent with early brainstem vulnerability, while indicating that descending cortical motor projections may be among the first large-scale pathways to show structural change.

In addition to these motor-system findings, we also identified reduced FC in the left inferior longitudinal fasciculus, a pathway linking occipital visual areas with limbic and anterior temporal regions. The greater severity of iRBD was associated with a reduction in the FC of the left inferior longitudinal fasciculus. Moreover, among RBD severity questionnaire subitems, the presence of vivid dreams was specifically linked to lower FC in the left inferior longitudinal fasciculus. This tract connects occipital visual areas with anterior temporal regions, including the amygdala and perirhinal cortex, that support object recognition, semantic, and affective processing of visual input.^58,60^ Notably, the affected left inferior longitudinal fasciculus, but not right, has been reported in Parkinson’s disease patients with visual hallucinations.^61–63^ Visual hallucinations have been associated with more frequent vivid dreams.^64^ Complementing these clinical stage disease findings, Li et al.^65^ showed that visual dysfunctions, particularly illusions rather than hallucinations, are nearly ten times more common in iRBD patients compared to healthy controls. The neurological basis of vivid dreams cannot be easily studied; however, De Gennaro et al.^66^ showed that higher dream vividness was linked with amygdala volumes in Parkinson’s disease, and the amygdala connects to higher-order visual cortices via an occipito-amygdalar segment of the inferior longitudinal fasciculus.^58^ Taken together, our findings indicate a lower FC of the left inferior longitudinal fasciculus correlating with more severe iRBD and vivid dreams, consistent with the possibility that early disruption of amygdalo-visual connectivity provides the anatomical substrate for progression from vivid dreaming and visual illusions to visual hallucinations that may later emerge. Notably, given the cross-sectional nature of our study, a longitudinal neuroimaging study will be necessary to confirm whether inferior longitudinal fasciculus damage precedes and predicts the onset of visual hallucinations.

Both the RBD severity questionnaire and polysomnography-based analyses highlighted the involvement of two major descending motor pathways, the corticospinal and frontopontine tracts, in prodromal Parkinson’s disease. Importantly, lower FD within the left corticospinal tract correlated with worse fine motor performance, underscoring its clinical relevance. The corticospinal tract is the principal tract for voluntary distal movements originating in the precentral gyrus,^67^ while the frontopontine tract carries premotor, executive, and limbic signals from the medial-frontal cortex to the pontine nuclei en route to the cerebellum; damage to either tract can disrupt movement execution or planning.^68^ Given its importance in movement, it is not surprising that alterations in the corticospinal tract have frequently been reported in Parkinson’s disease,^69–71^ and more recently in the prodromal cohorts.^12,72^ Our findings extend these observations by showing that corticospinal tract degeneration is selectively linked to iRBD severity (but not hyposmia) and already translates into fine motor loss.

Fine motor deficits refer to the inability to perform precise and small-amplitude actions, such as writing, buttoning a shirt, or picking up a coin. Such dexterity deficits are common in Parkinson’s disease patients and predict a higher risk of dementia.^73–76^ Dan et al.^77^ reported the presence of fine motor deficits in the unaffected body side, which was highly correlated with the time to convert to bilateral Parkinson’s disease. Fine motor deficits are not limited to clinical Parkinson’s disease. They are also reported in iRBD patients.^65,78,79^ Notably, the presence of fine motor deficits increases the likelihood of developing an α-synucleinopathy.^80^ These deficits are among the earliest motor symptoms appearing in Parkinson’s disease; poor performance on the Purdue pegboard test, a surrogate for dexterity, appears 7.5 years before the diagnosis of Parkinson’s disease.^3^ Neuroimaging studies remain limited; however, a recent population-based cohort study involving more than 8,000 participants found that lower fine motor performance was associated with a thinner precentral gyrus.^81^ In addition, lesions in the corticospinal tract in stroke patients result in severe fine motor control deficits in the ipsilateral paretic hand that, unlike other motor deficits, cannot be compensated by extrapyramidal pathways.^82^ Altogether, our findings indicate that corticospinal tract microstructure serves as an early marker of motor degeneration in prodromal Parkinson’s disease, which is reflected behaviorally in fine motor deficits.

We did not find any statistically significant volume- or diffusion-based correlates of hyposmia. Several biological or methodological factors may account for this null finding. First, olfactory dysfunction at this stage may stem more from synaptic alterations rather than gross axonal or grey matter loss.^83^ Second, according to the brain-first vs. body-first theory of Parkinson’s disease,^2^ synucleinopathy emerging in the olfactory bulb propagates through the smaller, short white matter bundles to nearby limbic structures (e.g., the amygdala).^84^ These small fiber bundles are poorly represented in standard fixel masks, reducing the power to detect fiber-specific changes, even though we observed a reduced fiber cross-section of the inferior longitudinal fasciculus, which connects temporal structures (including the amygdala) to occipital cortex, in association with more severe iRBD. Nonetheless, the possibility of including various causes of hyposmia (e.g., COVID-19) in a patient sample may result in a non-convergent finding, as each cause may have different neural correlates;^85^ however, the inclusion of reduced DaTscan as one of the criteria for the PPMI prodromal cohort makes this less likely. Future studies using larger sample sizes with olfactory bulb-specific imaging sequences seem necessary.

We did not detect any differences in grey matter volume associated with iRBD severity, UPSIT score, or polysomnography status. A plausible explanation is that the microstructural white matter changes precede the emergence of atrophy, a temporal sequence documented in neurodegenerative disorders such as Parkinson’s and Alzheimer’s diseases.^7,8^ Moreover, the structural connections provide a conduit along which misfolded α-synuclein spreads, giving the notion of “prion-like propagation” of synucleinopathies,^4,86^ implying that tract degeneration may precede and drive grey matter loss. Nonetheless, existing volumetric analysis literature on iRBD has shown volume loss in frontal and temporal regions compared to healthy controls,^87,88^ but these findings were based on case-control comparisons between patients and healthy controls. In contrast, our analysis examined symptom-structure associations within a larger mixed prodromal cohort (n = 88), in which grey matter effects are expected to be more subtle and spatially heterogeneous. Our larger within-group analysis did not replicate these earlier cortical findings, suggesting that grey matter changes in the prodromal phase may be less reliably detectable than fiber-specific microstructural alterations. Together, these results underscore that fixel-based diffusion metrics may provide a more sensitive marker of early neurodegenerative processes than conventional volumetric measures.

## 5. Conclusion

Our results show that FD and FC in the corticospinal, frontopontine, and inferior longitudinal fasciculi vary systematically with iRBD severity, indicating symptom-related white matter involvement during the prodromal phase. These associations emerged in the absence of corresponding grey matter relationships or broad motor impairments, suggesting that tract-specific microstructural changes may be more sensitive markers of prodromal pathology than volumetric alterations. The link between corticospinal fiber density and fine motor deficits further underscores the behavioural relevance of early disruption in this pathway. In summary, our fixel-based analyses indicate that descending motor tracts exhibit structural involvement that scales with iRBD severity, while the inferior longitudinal fasciculus demonstrates parallel visuolimbic involvement. Consistent corticospinal findings across questionnaire-based and polysomnography-confirmed iRBD point to early motor-pathway alterations as a potential hallmark of prodromal Parkinson’s disease. Longitudinal studies are needed to determine whether these tract-specific changes predict conversion to clinical Parkinson’s disease.

## Data Availability

The data is available on the PPMI website after agreeing to the dataset's data sharing policy (http://www.ppmi-info.org).

## Declarations

### Ethics approval and consent to participate

PPMI is a multi-site observational study conducted in accordance with the Declaration of Helsinki and Good Clinical Practice guidelines. The study protocol and informed consent documents were approved by the Institutional Review Board (IRB)/Ethics Committee at each participating site, and written informed consent was obtained from all participants (ClinicalTrials.gov identifier: NCT01141023).

### Consent for publication

-

## Competing Interests

The authors declare that they have no competing interests.

## Funding

This work was supported by the Research Training Group (RTG) 2783, funded by the German Research Foundation (DFG) - Project ID 456732630. SR was funded by the Deutsche Forschungsgemeinschaft (DFG, German Research Foundation; RO6114/2-1).

## Authors’ contributions

Conceptualization: A.A., S.R., M.T., C.M.T; Methodology: A.A., M.T., C.M.T.; Formal analysis: A.A.; Investigation: A.A.; Data curation: A.A.; Visualization: A.A.; Writing – original draft: A.A., S.R.; Writing – review & editing: A.A., S.R., M.T., C.M.T.; Supervision: C.M.T.; Project administration: A.A., C.M.T. All authors read and approved the final version of the manuscript.

## Acknowledgements

Data used in the preparation of this article were obtained in October 2024 from the Parkinson’s Progression Markers Initiative (PPMI) database (https://www.ppmi-info.org/access-data-specimens/download-data), RRID:SCR_006431. For up-to-date information on the study, visit http://www.ppmi-info.org.

PPMI – a public-private partnership – is funded by the Michael J. Fox Foundation for Parkinson’s Research and funding partners, including 4D Pharma, Abbvie, AcureX, Allergan, Amathus Therapeutics, Aligning Science Across Parkinson’s, AskBio, Avid Radiopharmaceuticals, BIAL, BioArctic, Biogen, Biohaven, BioLegend, BlueRock Therapeutics, Bristol-Myers Squibb, Calico Labs, Capsida Biotherapeutics, Celgene, Cerevel Therapeutics, Coave Therapeutics, DaCapo Brainscience, Denali, Edmond J. Safra Foundation, Eli Lilly, Gain Therapeutics, GE HealthCare, Genentech, GSK, Golub Capital, Handl Therapeutics, Insitro, Jazz Pharmaceuticals, Johnson & Johnson Innovative Medicine, Lundbeck, Merck, Meso Scale Discovery, Mission Therapeutics, Neurocrine Biosciences, Neuron23, Neuropore, Pfizer, Piramal, Prevail Therapeutics, Roche, Sanofi, Servier, Sun Pharma Advanced Research Company, Takeda, Teva, UCB, Vanqua Bio, Verily, Voyager Therapeutics, the Weston Family Foundation and Yumanity Therapeutics.

The analysis was performed at the high-performance cluster ROSA, located at the University of Oldenburg (Germany) and funded by the DFG through its Major Research Instrumentation Programme (INST 184/225-1 FUGG) and the Ministry of Science and Culture (MWK) of the Lower Saxony State.

## Notes

### Competing Interest Statement

The authors have declared no competing interest.

### Author Declarations

Data used in the preparation of this article were obtained in October 2024 from the Parkinsons Progression Markers Initiative (PPMI) database (https://www.ppmi-info.org/access-data-specimens/download-data), RRID:SCR_006431. For up-to-date information on the study, visit http://www.ppmi-info.org. PPMI is a multi-site observational study conducted in accordance with the Declaration of Helsinki and Good Clinical Practice guidelines. The study protocol and informed consent documents were approved by the Institutional Review Board (IRB)/Ethics Committee at each participating site, and written informed consent was obtained from all participants (ClinicalTrials.gov identifier: NCT01141023).

## References

1. Su, D. et al. Projections for prevalence of Parkinson’s disease and its driving factors in 195 countries and territories to 2050: modelling study of Global Burden of Disease Study 2021. BMJ 388, e080952 (2025).

2. Berg, D. et al. Prodromal Parkinson disease subtypes - key to understanding heterogeneity. Nat. Rev. Neurol. 17, 349–361 (2021).

3. Fereshtehnejad, S.-M. et al. Evolution of prodromal Parkinson’s disease and dementia with Lewy bodies: a prospective study. Brain 142, 2051–2067 (2019).

4. Steiner, J. A., Quansah, E. & Brundin, P. The concept of alpha-synuclein as a prion-like protein: ten years after. Cell Tissue Res. 373, 161–173 (2018).

5. Del Tredici, K. & Braak, H. Review: Sporadic Parkinson’s disease: development and distribution of α-synuclein pathology. Neuropathol. Appl. Neurobiol. 42, 33–50 (2016).

6. Ferreira, N. et al. Prodromal neuroinvasion of pathological α-synuclein in brainstem reticular nuclei and white matter lesions in a model of α-synucleinopathy. Brain Commun. 3, fcab104 (2021).

7. Jacobs, H. I. L. et al. Structural tract alterations predict downstream tau accumulation in amyloid-positive older individuals. Nat. Neurosci. 21, 424–431 (2018).

8. Rektor, I. et al. White matter alterations in Parkinson’s disease with normal cognition precede grey matter atrophy. PLoS One 13, e0187939 (2018).

9. Mitchell, T. et al. Emerging neuroimaging biomarkers across disease stage in Parkinson disease: A Review: A review. JAMA Neurol. 78, 1262–1272 (2021).

10. Ohlhauser, L., Smart, C. M. & Gawryluk, J. R. Tract-based spatial statistics reveal lower white matter integrity specific to idiopathic rapid eye movement Sleep Behavior Disorder as a proxy for prodromal Parkinson’s disease. J. Parkinsons. Dis. 9, 723–731 (2019).

11. Byun, J.-I. et al. White matter tract-specific microstructural disruption is associated with depressive symptoms in isolated RBD. NeuroImage Clin. 36, 103186 (2022).

12. Pimer, L. J. et al. Aberrant corticospinal tract characteristics in prodromal PD: A diffusion tensor imaging study. Clin. Park. Relat. Disord. 8, 100182 (2023).

13. Jeurissen, B., Leemans, A., Tournier, J.-D., Jones, D. K. & Sijbers, J. Investigating the prevalence of complex fiber configurations in white matter tissue with diffusion magnetic resonance imaging. Hum. Brain Mapp. 34, 2747–2766 (2013).

14. Raffelt, D. A. et al. Investigating white matter fibre density and morphology using fixel-based analysis. Neuroimage 144, 58–73 (2017).

15. Dhollander, T. et al. Fixel-based Analysis of diffusion MRI: Methods, applications, challenges and opportunities. Neuroimage 241, 118417 (2021).

16. Rau, Y.-A. et al. A longitudinal fixel-based analysis of white matter alterations in patients with Parkinson’s disease. NeuroImage Clin. 24, 102098 (2019).

17. Zarkali, A., McColgan, P., Leyland, L.-A., Lees, A. J. & Weil, R. S. Visual dysfunction predicts cognitive impairment and white matter degeneration in Parkinson’s disease. Mov. Disord. 36, 1191–1202 (2021).

18. Zarkali, A. et al. Neuroimaging and plasma evidence of early white matter loss in Parkinson’s disease with poor outcomes. Brain Commun. 6, fcae130 (2024).

19. Zhou, W. et al. Fiber-specific white matter alterations in Parkinson’s disease patients with freezing of gait. Brain Res. 1815, 148440 (2023).

20. Marek, K. et al. The Parkinson’s progression markers initiative (PPMI) – establishing a PD biomarker cohort. Ann. Clin. Transl. Neurol. 5, 1460–1477 (2018).

21. Stiasny-Kolster, K. et al. The REM sleep behavior disorder screening questionnaire--a new diagnostic instrument. Mov. Disord. 22, 2386–2393 (2007).

22. Doty, R. L., Shaman, P. & Dann, M. Development of the University of Pennsylvania Smell Identification Test: a standardized microencapsulated test of olfactory function. Physiol. Behav. 32, 489–502 (1984).

23. Goetz, C. G. et al. Movement Disorder Society□sponsored revision of the Unified Parkinson’s Disease Rating Scale (MDS□UPDRS): Scale presentation and clinimetric testing results. Mov. Disord. 23, 2129–2170 (2008).

24. Cella, D. et al. Neuro-QOL: brief measures of health-related quality of life for clinical research in neurology. Neurology 78, 1860–1867 (2012).

25. Tournier, J.-D. et al. MRtrix3: A fast, flexible and open software framework for medical image processing and visualisation. Neuroimage 202, 116137 (2019).

26. Veraart, J. et al. Denoising of diffusion MRI using random matrix theory. Neuroimage 142, 394–406 (2016).

27. Kellner, E., Dhital, B., Kiselev, V. G. & Reisert, M. Gibbs-ringing artifact removal based on local subvoxel-shifts. Magn. Reson. Med. 76, 1574–1581 (2016).

28. Andersson, J. L. R. & Sotiropoulos, S. N. An integrated approach to correction for off-resonance effects and subject movement in diffusion MR imaging. Neuroimage 125, 1063–1078 (2016).

29. Andersson, J. L. R., Skare, S. & Ashburner, J. How to correct susceptibility distortions in spin-echo echo-planar images: application to diffusion tensor imaging. Neuroimage 20, 870–888 (2003).

30. Smith, S. M. et al. Advances in functional and structural MR image analysis and implementation as FSL. Neuroimage 23 **Suppl 1**, S208–19 (2004).

31. Andersson, J. L. R., Graham, M. S., Zsoldos, E. & Sotiropoulos, S. N. Incorporating outlier detection and replacement into a non-parametric framework for movement and distortion correction of diffusion MR images. Neuroimage 141, 556–572 (2016).

32. Andersson, J. L. R. et al. Towards a comprehensive framework for movement and distortion correction of diffusion MR images: Within volume movement. Neuroimage 152, 450–466 (2017).

33. Tustison, N. J. et al. N4ITK: Improved N3 Bias Correction. IEEE Trans. Med. Imaging 29, 1310–1320 (2010).

34. Genc, S. et al. Impact of b-value on estimates of apparent fibre density. Hum. Brain Mapp. 41, 2583–2595 (2020).

35. Dhollander, T., Raffelt, D. & Connelly, A. Unsupervised 3-tissue response function estimation from single-shell or multi-shell diffusion MR data without a co-registered T1 image. in 5 (2016).

36. Dhollander, T., Mito, R., Raffelt, D. & Connelly, A. Improved white matter response function estimation for 3-tissue constrained spherical deconvolution. in *Proc*. Intl. Soc. Mag. Reson. Med vol. 555 (archive.ismrm.org, 2019).

37. Billot, B. et al. SynthSeg: Segmentation of brain MRI scans of any contrast and resolution without retraining. Med Image Anal 86, 102789 (2023).

38. Billot, B. et al. Robust machine learning segmentation for large-scale analysis of heterogeneous clinical brain MRI datasets. Proc Natl Acad Sci U S A 120, e2216399120 (2023).

39. Dhollander, T. & Connelly, A. A novel iterative approach to reap the benefits of multi-tissue CSD from just single-shell (+b=0) diffusion MRI data. in *Proc*. Intl. Soc. Mag. Reson. Med. (2016).

40. Raffelt, D. et al. Bias Field Correction and Intensity Normalisation for Quantitative Analysis of Apparent Fibre Density. in Proc. Intl. Soc. Mag. Reson. Med.

41. Dhollander, T., et al. Multi-tissue log-domain intensity and inhomogeneity normalisation for quantitative apparent fibre density. in Proc. Intl. Soc. Mag. Reson. Med. (2021).

42. Smith, R. E., Tournier, J.-D., Calamante, F. & Connelly, A. SIFT: Spherical-deconvolution informed filtering of tractograms. Neuroimage 67, 298–312 (2013).

43. Raffelt, D. et al. Apparent Fibre Density: a novel measure for the analysis of diffusion-weighted magnetic resonance images. Neuroimage 59, 3976–3994 (2012).

44. Tournier, J., Calamante, F. & Connelly, A. Improved probabilistic streamlines tractography by 2 nd order integration over fibre orientation distributions. in Proceedings of the International Society for Magnetic Resonance in Medicine (2010).

45. Wasserthal, J., Neher, P. & Maier-Hein, K. H. TractSeg - Fast and accurate white matter tract segmentation. Neuroimage 183, 239–253 (2018).

46. Wasserthal, J., Neher, P. F., Hirjak, D. & Maier-Hein, K. H. Combined tract segmentation and orientation mapping for bundle-specific tractography. Med. Image Anal. 58, 101559 (2019).

47. Wasserthal, J., Neher, P. F. & Maier-Hein, K. H. Tract orientation mapping for bundle-specific tractography. arXiv [cs.CV] (2018).

48. Posit Team. RStudio: Integrated Development Environment for R. (2023).

49. R Core Team. R: A Language and Environment for Statistical Computing. (2023).

50. Jackman, S. pscl: Classes and Methods for R Developed in the Political Science Computational Laboratory. Preprint at https://github.com/atahk/pscl/ (2024).

51. Zeileis, A., Kleiber, C. & Jackman, S. Regression Models for Count Data in R. Journal of Statistical Software vol. 27 Preprint at https://www.jstatsoft.org/v27/i08/ (2008).

52. Mullahy, J. Specification and testing of some modified count data models. J. Econom. 33, 341–365 (1986).

53. Feng, C. X. A comparison of zero-inflated and hurdle models for modeling zero-inflated count data. J. Stat. Distrib. Appl. 8, 8 (2021).

54. Benjamini, Y. & Hochberg, Y. Controlling the false discovery rate: A practical and powerful approach to multiple testing. J. R. Stat. Soc. Series B Stat. Methodol. 57, 289–300 (1995).

55. Gaser, C. et al. CAT: a computational anatomy toolbox for the analysis of structural MRI data. Gigascience 13, giae049 (2024).

56. Ashburner, J. & Friston, K. J. Unified segmentation. Neuroimage 26, 839–851 (2005).

57. Klein, A. et al. Evaluation of 14 nonlinear deformation algorithms applied to human brain MRI registration. Neuroimage 46, 786–802 (2009).

58. Latini, F. New insights in the limbic modulation of visual inputs: the role of the inferior longitudinal fasciculus and the Li-Am bundle. Neurosurg. Rev. 38, 179–89; discussion 189–90 (2015).

59. Zhou, S. et al. Brain structural basis of individual variability in dream recall frequency. Brain Imaging Behav. 13, 1474–1485 (2019).

60. Herbet, G., Zemmoura, I. & Duffau, H. Functional anatomy of the inferior longitudinal fasciculus: From historical reports to current hypotheses. Front. Neuroanat. 12, 77 (2018).

61. Yuki, N., Yoshioka, A., Mizuhara, R. & Kimura, T. Visual hallucinations and inferior longitudinal fasciculus in Parkinson’s disease. Brain Behav. 10, e01883 (2020).

62. Zarkali, A. et al. Fiber-specific white matter reductions in Parkinson hallucinations and visual dysfunction. Neurology 94, e1525–e1538 (2020).

63. Lenka, A. et al. Abnormalities in the white matter tracts in patients with Parkinson disease and psychosis. Neurology 94, e1876–e1884 (2020).

64. Gama, R. L. et al. Risk factors for visual hallucinations in patients with Parkinson’s disease. Neurol. Res. 37, 112–116 (2015).

65. Li, Y. et al. Visual dysfunction in patients with idiopathic rapid eye movement sleep behavior disorder. Neurosci. Lett. 709, 134360 (2019).

66. De Gennaro, L., et al. Dopaminergic system and dream recall: An MRI study in Parkinson’s disease patients: Dopaminergic System and Dreaming. Hum. Brain Mapp. 37, 1136–1147 (2016).

67. Welniarz, Q., Dusart, I. & Roze, E. The corticospinal tract: Evolution, development, and human disorders. Dev. Neurobiol. 77, 810–829 (2017).

68. Schmahmann, J. D. & Pandya, D. N. Anatomic organization of the basilar pontine projections from prefrontal cortices in rhesus monkey. J. Neurosci. 17, 438–458 (1997).

69. Andica, C. et al. Fiber-specific white matter alterations in early-stage tremor-dominant Parkinson’s disease. NPJ Parkinsons Dis. 7, 51 (2021).

70. Chen, N.-K. et al. Alteration of diffusion-tensor magnetic resonance imaging measures in brain regions involved in early stages of Parkinson’s disease. Brain Connect. 8, 343–349 (2018).

71. Atkinson-Clement, C., Pinto, S., Eusebio, A. & Coulon, O. Diffusion tensor imaging in Parkinson’s disease: Review and meta-analysis. NeuroImage Clin. 16, 98–110 (2017).

72. Campabadal, A., Segura, B., Junque, C. & Iranzo, A. Structural and functional magnetic resonance imaging in isolated REM sleep behavior disorder: A systematic review of studies using neuroimaging software. Sleep Med. Rev. 59, 101495 (2021).

73. Zhang, J. et al. Comprehensive assessment of fine motor movement and cognitive function among older adults in China: a cross-sectional study. BMC Geriatr. 24, 118 (2024).

74. Teulings, H. L., Contreras-Vidal, J. L., Stelmach, G. E. & Adler, C. H. Parkinsonism reduces coordination of fingers, wrist, and arm in fine motor control. Exp. Neurol. 146, 159–170 (1997).

75. Dahdal, P. et al. Fine motor function skills in patients with Parkinson disease with and without mild cognitive impairment. Dement. Geriatr. Cogn. Disord. 42, 127–134 (2016).

76. Iakovakis, D. et al. Touchscreen typing-pattern analysis for detecting fine motor skills decline in early-stage Parkinson’s disease. Sci. Rep. 8, 7663 (2018).

77. Dan, X. et al. Impaired fine motor function of the asymptomatic hand in unilateral Parkinson’s disease. Front. Aging Neurosci. 11, 266 (2019).

78. Elliott, J. E. et al. Baseline characteristics of the North American prodromal Synucleinopathy cohort. Ann. Clin. Transl. Neurol. 10, 520–535 (2023).

79. Maetzler, W. & Hausdorff, J. M. Motor signs in the prodromal phase of Parkinson’s disease. Mov. Disord. 27, 627–633 (2012).

80. Postuma, R. B. et al. Risk and predictors of dementia and parkinsonism in idiopathic REM sleep behaviour disorder: a multicentre study. Brain 142, 744–759 (2019).

81. Yang, X., Zeng, W., Estrada, S., Breteler, M. M. B. & Aziz, N. A. Association between brain structure and fine motor function: findings from the population-based Rhineland Study. EBioMedicine 116, 105771 (2025).

82. Paul, T. et al. The role of corticospinal and extrapyramidal pathways in motor impairment after stroke. Brain Commun. 5, fcac301 (2023).

83. Chen, F. et al. α-Synuclein aggregation in the olfactory bulb induces olfactory deficits by perturbing granule cells and granular-mitral synaptic transmission. NPJ Parkinsons Dis. 7, 114 (2021).

84. Ubeda-Bañon, I., Saiz-Sanchez, D., de la Rosa-Prieto, C. & Martinez-Marcos, A. α-Synuclein in the olfactory system in Parkinson’s disease: role of neural connections on spreading pathology. Brain Struct. Funct. 219, 1513–1526 (2014).

85. Yildirim, D., Kandemirli, S. G., Tekcan Sanli, D. E., Akinci, O. & Altundag, A. A comparative olfactory MRI, DTI and fMRI study of COVID-19 related anosmia and post viral olfactory dysfunction. Acad. Radiol. 29, 31–41 (2022).

86. Jan, A., Gonçalves, N. P., Vaegter, C. B., Jensen, P. H. & Ferreira, N. The prion-like spreading of alpha-synuclein in Parkinson’s disease: Update on models and hypotheses. Int. J. Mol. Sci. 22, 8338 (2021).

87. Rahayel, S. et al. Abnormal gray matter shape, thickness, and volume in the motor Cortico-subcortical loop in idiopathic rapid eye movement sleep behavior disorder: Association with clinical and motor features. Cereb. Cortex 28, 658–671 (2018).

88. Pereira, J. B. et al. Cortical thinning in patients with REM sleep behavior disorder is associated with clinical progression. NPJ Parkinsons Dis. 5, 7 (2019).

