## Supplementary material for "Early degeneration of motor pathways in prodromal Parkinson’s disease: A fixel-based structural connectivity analysis"

**Supplementary Table 1.** Scales from MDS-UPDRS part 3 questionnaire used to calculate subscale scores of rigidity, bradykinesia, and rest tremor.

**Supplementary Table 2.** Fixel overlaps with the tracts generated by TractSeg.

**Supplementary Figure 1.** Histograms of motor assessments in our sample.

**Supplementary Figure 2.** Higher severity of RBD is associated with lower fiber cross-section in the left temporal white matter, right internal capsule, and brainstem.

**Supplementary Figure 3.** Voxel-based morphometry analysis results.

**Supplementary Table 1.** Scales from MDS-UPDRS part 3 questionnaire that were used to calculate subscale scores of rigidity, bradykinesia, and rest tremor. The subscale score was calculated as the sum of scales. RUE: right upper extremity, LUE: left upper extremity, RLE: right lower extremity, LLE: left lower extremity.

| **Subscale** | **Scales** |
|---|---|
| Rigidity | Rigidity (3.3): Neck, RUE, LUE, RLE, LLE |
| Bradykinesia | Finger Tapping (3.4): right and left<br>Hand Movements (3.5): right and left<br>Pronation-Supination Movements of Hands (3.6): right and left<br>Toe Tapping (3.7): right and left<br>Leg Agility (3.8): right and left |
| Rest Tremor | Rest Tremor Amplitude (3.17): RUE, LUE, RLE, LLE, Lip/Jaw<br>Constancy of Rest Tremor (3.18) |

**Supplementary Table 2.** The number of significant fixels overlapping with the tracts generated by TractSeg sorted in descending order.

| Tract | Number of overlaps |
|---|---|
| Right corticospinal tract | 27 |
| Right parieto-occipito-pontine tract | 27 |
| Right frontopontine tract | 18 |
| Left Inferior longitudinal tract | 13 |
| Left arcuate fasciculus | 13 |
| Middle longitudinal fasciculus | 13 |
| Right superior cerebellar peduncle | 12 |
| Right superior thalamic radiation | 12 |
| Right thalamoparietal | 12 |
| Right thalamo-postcentral | 12 |
| Right thalamo-precentral | 12 |
| Left inferior fronto-occipital | 4 |
| Right striatoparietal | 3 |
| Right striatoprecentral | 3 |

**Supplementary Figure 1.** Histograms of motor assessments in our sample. The prodromal subjects do not manifest clinical Parkinson's disease symptoms, which explains the negative binomial distribution of motor symptoms in our sample.

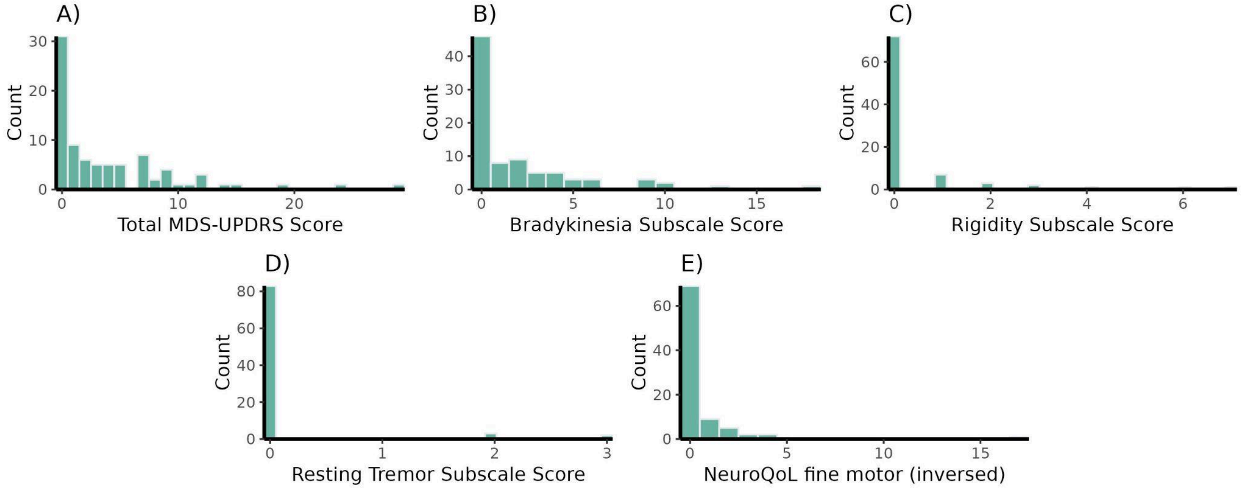

**Supplementary Figure 2. Higher severity of RBD is associated with lower fiber cross-section in the left temporal white matter, right internal capsule, and brainstem.** The fixels are color-coded; blue-purple and green show fibers stretched along superior-inferior, and anterior-posterior axes, respectively.

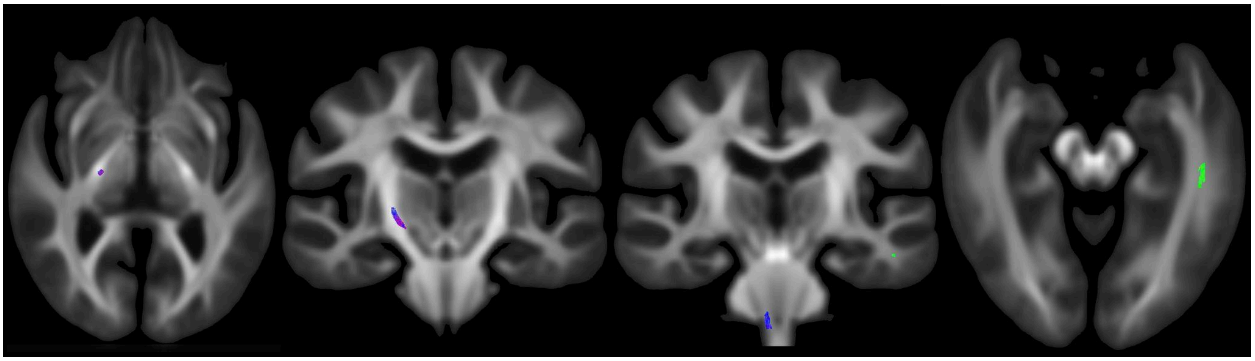

**Supplementary Figure 3. Voxel-based morphometry analysis results.** A cluster of voxels showing reduced grey matter volume in polysomnography-positive REM sleep behavior disorder compared to the subjects with negative polysomnography findings. The peak shown was located in MNI coordinates of 32 -4 -44, mapped to the right temporal fusiform gyrus, according to the Harvard-Oxford atlas (uncorrected $p = 0.007$, family-wise error-corrected $p = 0.079$).

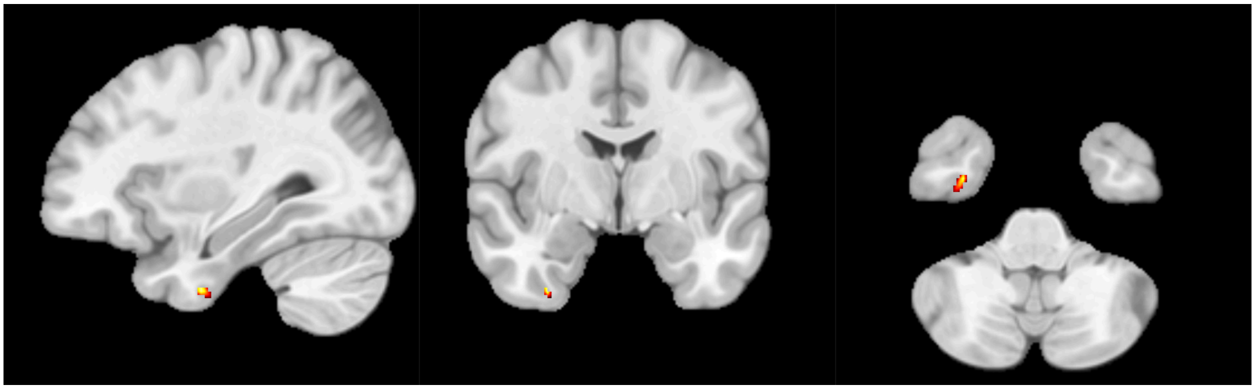
